# Incidence and prognostic factors of knee extension deficits following anterior cruciate ligament reconstruction: A systematic review and meta-analysis of randomised controlled trials

**DOI:** 10.1101/2020.11.26.20239046

**Authors:** Corey Scholes, Nalan Ektas, Meredith Harrison-Brown, Maha Jegatheesan, Ashwin Rajesh, Garry Kirwan, Christopher Bell

## Abstract

**Background and aims:** Knee extension deficits complicate recovery from ACL injury and reconstruction, however the incidence of knee extension loss is not well defined. The aim of this review was to identify the incidence of loss of extension (LOE) following ACL rupture and reconstruction, explore the definitions of knee extension deficits reported and identify prognostic factors affecting LOE incidence.

**Methods and analysis:** A systematic search was conducted in Medline, Cochrane Library and PEDro for studies in publication up to November 2021, with no restrictions on publication year. References were screened and assessed for inclusion using predetermined eligibility criteria. Randomised controlled trials (RCTs) that quantified knee angle, loss of extension or incidence of extension deficit were included for quality assessment and data extraction. Statistical summaries were generated and meta-analyses performed in two parts to examine: (i) the probability of a datapoint being zero incidence compared to a non-zero incidence, and (ii) the relationship between the predictors and non-zero LOE incidence.

**Results:** A sample of 15494 papers were retrieved using the search criteria, with 53 studies meeting eligibility criteria. Pooled results from 4991 participants were included for analysis, with 4891 participants who had undergone ACLR. The proportion of included studies judged at an overall low risk of bias was small (7.8%). The observed group and study were the most important predictors for whether a datapoint reported an incidence of extension deficit. Time to follow up (P < 0.001) and graft type (P = 0.02) were found to have a significant influence on non-zero LOE incidence (%). Covariate adjusted estimates of average LOE indicated 1 in 3 patients presenting with LOE at 12months followup, reducing to 1 in 4 at 2 years.

**Conclusions:** This review examined the definitions for the measurement and interpretation of postoperative knee extension, and established the trajectory of knee extension deficit after ACL injury and reconstruction. While factors associated with loss of extension were identified, the exact trajectory of knee extension deficits were difficult to infer due to discrepancies in measurement techniques and patient variation. Clinicians should expect up to 1 in 3 patients to present postoperatively with loss of extension of at least 3 degrees, which may resolve in some patients over time. Future work should focus on LOE as a clinically relevant complication of ACL injury and treatment with appropriate attention to standardisation of definitions, measurements and better understanding of natural history.

**PROSPERO registration number:** CRD42018092295

## INTRODUCTION

Anterior cruciate ligament (ACL) rupture can have a long-lasting negative impact on a person’s health and quality of life. Treatments for ACL rupture comprise conservative non-operative management (such as activity modification, bracing or physical and activity based therapies), surgical repair, biological therapies or reconstruction of the ligament offered as a first-line treatment in some patients or following failed nonoperative management in others^1^. ACL reconstruction (ACLR) is a common treatment strategy employed to limit the long term complications following ACL rupture. Patients who undergo ACLR present with fewer subsequent meniscal injuries, lower incidence of subsequent surgery, and significantly improved activity levels compared to patients receiving non-operative management of their ACL rupture^2^ and with promising return to sport or activity rates observed in those who have had the procedure^3^.

Complications following ACL reconstruction (ACLR), though rare, can include missed concomitant injuries, tunnel malposition, infection, tunnel osteolysis, fixation failure, fracture, increased knee stiffness, graft site morbidity, and thromboembolic events^4,5^. Knee stiffness, presenting as either loss of extension and/or loss of flexion, is a common presentation following acute ACL injury and reconstruction, resulting in poorer functional outcomes and greater incidence of osteoarthritis^6^. Loss of knee extension is less tolerated than flexion loss, and contributes to limitations in athletic performance, functional deficits and increased risk of patellofemoral arthritis at extension deficits of 5 degrees or more^6,7^. The aetiology for loss of extension (LOE) is multifactorial, ranging from anterior-intercondylar notch scar tissue or graft impingement, capsulitis, technicalities of the surgery and choice of rehabilitation protocol ^7^.

Extension deficits remain a common cause of failure and revision surgery following primary ACLR^8^. While knee extension loss may be resolved with additional interventions, such as arthroscopy, manipulation under anaesthesia, pharmacologic(al) or physical therapies^9^, there is limited evidence available that identifies the point at which extension loss becomes detrimental to an individual’s functional recovery. While the degree of extension at early follow-up (4 weeks) was determined to be strongly coupled with the incidence at later phases of recovery (12 weeks follow-up)^10^, the true incidence of deficits are difficult to ascertain due to a lack of consistent criteria and poor definitions for determining loss of knee range of motion (ROM) across the literature^9^. The reported incidence of knee stiffness after surgical reconstruction of the ACL ranges from 2-35%^6^, with one retrospective study of 100 patients reporting the incidence of knee stiffness at 12% 6 months following ACL reconstruction ^11^. Specific incidence of loss of extension is also varied. A recent study of 229 patients reported 25.3% incidence at four weeks follow up^12^, while reviews of historical ACLR literature^7^ report up to 59% incidence of extension loss following ACLR. However extension loss is not commonly reported as a primary outcome for investigation across the literature. As such, the natural trajectory of knee extension following ACL rupture and true incidence of extension deficits after surgical reconstruction remains largely unknown, limiting the ability to develop standardised benchmarks for postoperative monitoring of knee extension recovery after ACL reconstruction.

### Objectives

The primary objective of this review was to identify the incidence of loss of knee extension angle in patients diagnosed with ACL rupture electing to undergo formal treatment under the care of a registered clinical provider (including non-operative and arthroscopic ACL reconstruction) compared to the contralateral limb or control patients, as reported within randomised control studies. The secondary objectives were to explore the definitions of knee extension deficits reported and the factors affecting the incidence of extension deficits.

## METHODS

This review was reported as per the Preferred Reporting Items for Systematic Review and Meta-analysis (PRISMA) statement^13^. A protocol for the systematic review was established a priori^14^ and registered on the PROSPERO International Prospective Register of Systematic Reviews, registration number CRD42018092295.

### Eligibility criteria

The PICOS (Population, Intervention, Comparison, Outcomes, Study Design) framework was used to formulate the research question and define eligibility criteria (Table A) for the literature search^15^.

**Table A:** Eligibility criteria for study inclusion

| Criteria | Inclusion | Exclusion |
| --- | --- | --- |
| **Population** | Adults diagnosed with ACL rupture.<br>Exclusions for patient sex, age or activity level were not applied. | Participants diagnosed with multiple-ligament rupture or patellar dislocation secondary to ACL rupture. |
| **Intervention** | Formal interventions used in the standard care and management of ACL rupture, including:<br>• Arthroscopic reconstruction of the ACL using autograft or allograft with any preparation and independent drilling of the femur and tibia and anatomical graft placement (footprint centre to footprint centre), and<br>• Conservative management using<br>○ Surgical repair of the ligament or<br>○ Injectable therapies (e.g. platelet-rich-plasma)<br>○ Rehabilitation or physical exercise therapy | Revision ACLR procedure, or ACL reconstruction or repair associated with joint preserving surgeries for unicompartmental degenerative disease treated with:<br>• Tibial or femoral osteotomy<br>• Meniscus transplantation<br>• Meniscal prosthesis implantation<br>• Joint arthroplasty. |
| **Comparators** | Minimum knee extension angle or extension deficit reported in the contralateral non-affected limb or in individuals unaffected by ACL rupture (identified as “healthy controls”) |  |
| **Outcomes** | Studies examining or reporting:<br>1. Knee extension angle (°) - in studies that have conducted measurements while patients are seated, prone or supine without muscle contraction to achieve minimum knee flexion.<br>2. Knee extension angle (°) - studies that have conducted measurements of knee angle during an active/locomotive task.<br>3. Incidence of knee extension deficit (%) - when the index knee is compared to the contralateral knee.<br>4. Incidence of knee extension loss (%) in longitudinal, repeated measures studies that have compared post intervention knee extension to the pre-intervention value for the same patients. | Studies that did not report variability measures around the extension angle or loss of extension angle were excluded. |
| **Study Design** | Randomised controlled trials. No restrictions on publication year were applied.<br>N.B. Systematic reviews identified during the search strategy were used to source additional primary materials but were not included in the analysis. The results of meta-analyses were included as a study in the analysis if they meet aforementioned inclusion criteria. | Studies not published in english language. |

### Information Sources

A comprehensive literature search of the following electronic databases was undertaken from their date of inception until September 2019: Pubmed for Medline, Cochrane Library from EBM Reviews (complete via Ovid SP) and Physiotherapy Evidence Database (PEDro). The search terms comprised keyword combinations and medical subject headings (MeSH) relating to three key domains identifying the pathology (ACL tear OR rupture), intervention or treatment strategy (conservative or surgical reconstruction) and outcomes of interest (extension deficit, loss of extension, stiffness, arthrofibrosis or range of motion). A comprehensive search strategy is presented in **Supplementary Material A - Study Search Strategy**.

### Study identification, selection and data extraction

The PRISMA flow process for systematic reviews ^13^ was used to identify, screen, and assess the eligibility and inclusion of studies to the review. Citations identified during the initial search were uploaded to a web-based bibliographic software (Paperpile LLC, Vienna, Austria) for study management and removal of duplicate records. Study identification was performed iteratively, with reference lists of identified systematic reviews searched to identify further references and retrieved from Medline using an application programming interface ^16^. Reference lists of eligible studies were also retrieved to identify additional studies for screening (Figure 1). A custom algorithm was coded to conduct a keyword search within identified titles based on inclusion and exclusion criteria to automatically identify titles for abstract screening, flag for manual title screening or exclude from further screening (Table B). Study screening for inclusion of studies to the review and data extraction was performed independently by a core team (BLL, CS, GK, NE, MJ, MHB) with discrepancies resolved with discussion.

**Table B:** Logic rules for custom algorithms coded for preliminary title screening during systematic review.

| Inclusion keyword | Logic | Exclusion keyword | Action |
| --- | --- | --- | --- |
| Multiple inclusion keywords found in title | AND | Exclusion keyword not found in title | Flag for abstract screen |
| Inclusion keywords not found in title | AND / OR | Exclusion keyword found in title | Flag as excluded |
| Inclusion keyword/s found in title | AND | Exclusion keyword not found in title | Flag for manual title screen |

**Figure 1:**
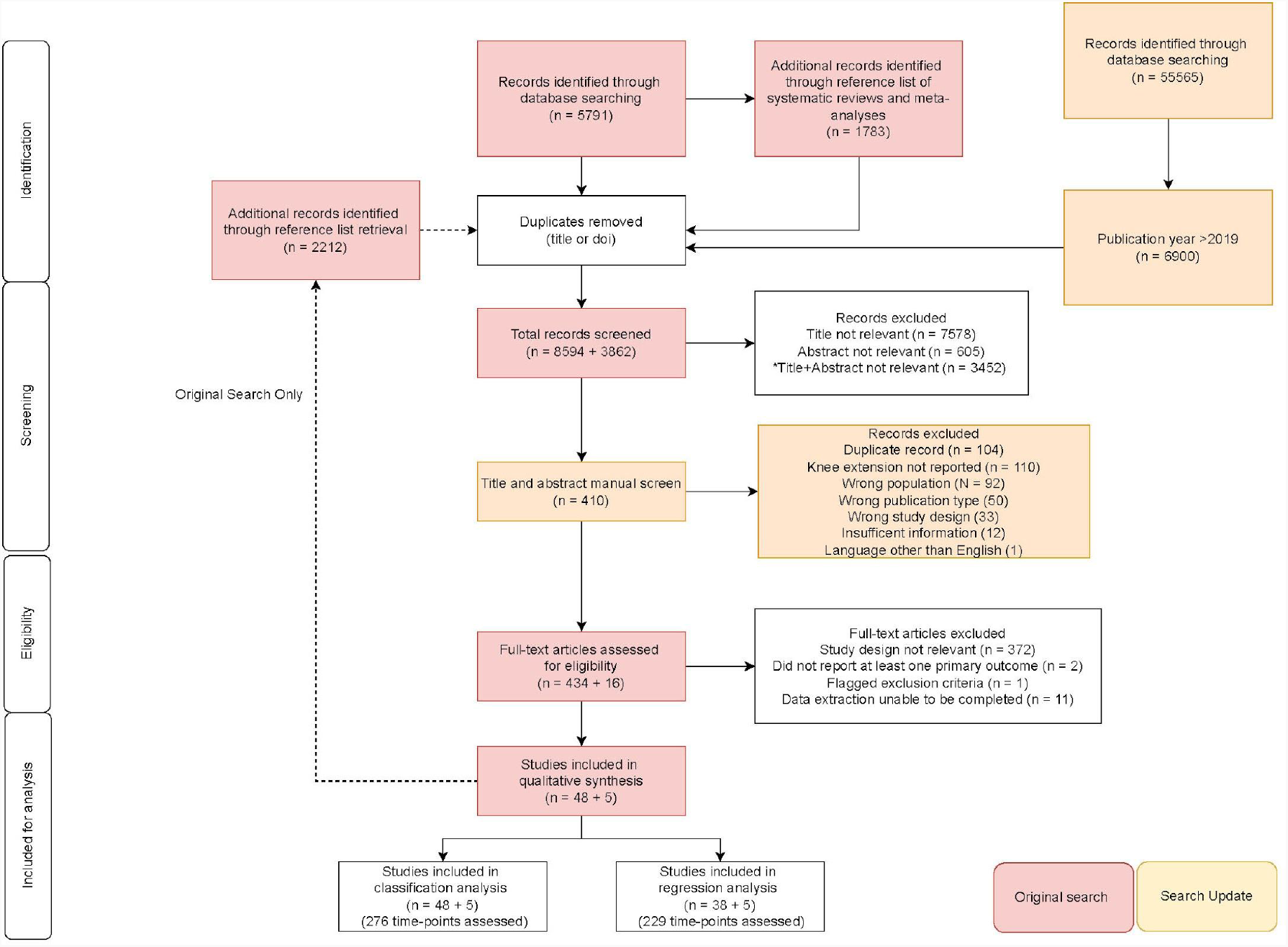
PRISMA flow diagram for study inclusion and data extraction for analysis.

Study and population characteristics, as well as intervention or surgical data and outcomes were extracted from included studies for investigation of the primary and secondary objectives of the review (Table C). Web-based forms were used (G Suite, Google, California, USA) for data extraction over three tiers: study, group, and time-point, with distinct variables and outcomes captured at each level depending on whether the data was unique to the study, an observed group within a study or an observed time-point. Screening and extracted data was stored on a cloud-based database (G Suite, Google, California, USA) and summarised to perform statistical analysis and meta-analysis where deemed eligible.

**Table C:** Data items extracted for the review.

| Study items | Demographic items | Intervention / treatment items | Outcomes |
| --- | --- | --- | --- |
| First author | Number of participants | Intervention mode | Time-point for data collection |
| Study title | Gender (% female) |  | Average knee extension angle |
| Year | Average age at treatment | For ACLR: |  |
| Study design | Average height | Fixation type | Average loss of knee extension angle (deg) |
| Number of unique groups | Average weight | Graft type | Knee extension loss incidence (%) |
| Comparator | Concomitant injuries | Graft tissue |  |
| Knee extension threshold | Comorbidities | Graft preparation | Significance of findings |
| Definition of extension deficit / LOE | Average time to intervention | Bundle |  |
| Measurement method | Average follow up period | Graft placement - approach and strategy |  |
| Movement paradigm |  | Whether notchplasty was performed |  |

### Search Update

An updated search was performed in November 2021. Searches were performed with the terms in the protoco ^14^ in post-review databases (Pubmed, Cochrane Trials Database and The Lens (lens.org)). The Lens is an open-access platform archiving hundreds of millions of scholarly articles. Search results were exported to ris format and combined using custom code (RStudio, v2021.09.0, build 351, RStudio PBC) drawing on the following packages (Synthesisr, Data.table, Tidyverse, Expss, Revtools ^17^, dplyr, stringi ^18^), to filter by publication year (>2019); remove exact and fuzzy duplicate matches of title and doi; remove systematic reviews and meta-analyses and then identify relevant articles by abstract keyword search **(Supplementary Material B - Combine Screening Code)**. The screened results were imported into Rayyan ^19^ for manual screening (Figure 1).

### Risk of bias

Independent scoring of risk of bias for included studies was performed by a third observer using the Revised Cochrane risk-of-bias tool for randomized trials; RoB2 ^20^. The RoB2 comprises grading the risk of bias across five distinct domains: randomisation process, deviations from intended interventions, missing outcome data, measurement of the outcome, and selection of the reported result. A single observer (MHB) performed the risk of bias assessment relative to the outcome of interest (loss of knee extension angle) using the Excel tool available at riskofbias.info ^20^. Each domain was rated a judgement of “low”, “some concerns” or “high” risk of bias based on the answers to the signalling questions. Domain ratings using the algorithm were overridden if considered inaccurate relative to the type of study. Overall risk of bias for included studies was scored “Low risk” where the study was judged to be at a low risk of bias across all domains; overall risk of bias was scored as “Some concerns” where the assessor raised some concern in at least one domain, but not at “High risk” for any domain; studies were judged at overall “High risk” where a high risk was captured in at least one domain, or some concerns were noted in multiple domains ^20^.

### Statistical analysis

Continuous variables were summarised using median and interquartile ranges, categorical variables were summarised as percentages (%). Primary outcomes, knee extension angle and loss of knee extension, were re-coded such that full extension was defined as zero degrees of flexion. The incidence of LOE was estimated from studies that reported an average angle (mean or median) and variance measures (interquartile range or standard deviation), using a normal distribution function (Equation 1)(*normdist* function, Sheets, Google Inc. USA), with the summary statistics (average knee angle (AngleAve) and standard deviation (AngleSD) and a threshold of 3 degrees as inputs.

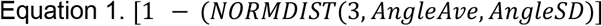

Knee extension angles were estimated for studies reporting heel height differences (HHD) using a previously described conversion ^21^.To identify factors that may be associated with the reported LOE incidence and estimate an adjusted LOE incidence, the analysis was conducted in two parts. The first part examined the probability of a datapoint being zero incidence compared to a non-zero incidence (classification analysis). The LOE incidence was re-coded to a binary categorical variable (detected; not detected). The predictor set was manually identified from the data items extracted. The main criteria for inclusion of a predictor was i) sufficient variation in responses; and ii) sufficient coverage in reporting (low rate of missingness). A feature reduction technique based on mutual information (MI) criterion was implemented to reduce the number of categorical predictors ^22^. The slope of the drop in MI was used to determine the inclusion of categorical variables, which were combined with continuous data predictors for the classification model. A bootstrap-aggregated decision tree^23^ was applied to the binary LOE categorisation (LOE 0; LOE>0) using the reduced predictor set via a treebagger model with 5-fold cross-validation. The model error was retrieved as an indicator of model fit and predictors were ranked based on the classification error attributed to each predictor. The second part of the metaregression provided an adjusted estimate of LOE by firstly subsetting the data based on non-zero incidence of LOE and assessed the relationship between the predictors and LOE incidence. A weighted (by study sample size) generalised mixed-effects linear model (normal distribution, Laplace fit method) was applied. Fixed effects incorporated study methodology, surgical technique, followup timing and risk of bias. Random effects were group identifiers nested within studies. Overall model fit was assessed with adjusted R^2^. Fixed effects estimates (with confidence intervals) and p-values were summarised. The analyses were conducted within a dedicated statistics toolbox of Matlab (v9.7 (2020b), Mathworks Inc, USA).

## RESULTS

### Study selection and characteristics

The search results (including *updated* search) combined with citation retrieval returned a total of 15494 non-duplicate records for title screening. A sample of 53 randomised trials ^24–77^ met the inclusion criteria and were examined (Figure 1). The included papers defined 113 patient groups with 276 instances of LOE (timepoints). The studies included were published between 1987 and 2021 and study groups were randomised by ACL graft type (43.4%), knee range of motion restriction such as use of continuous passive motion, bracing and other interventions (20.8%), surgical technique (15.1% - graft placement and fixation, adjunct procedures and the ACL remnant), the timing of interventions (9.4% - ACL reconstruction and bracing) and rehabilitation (11.3%). A summary of studies is available in **Supplementary Material C - Summary of Studies**.

The proportion of included studies judged at an overall low risk of bias was small (N = 4, 7.6%), with 41.5% and 50.9% of studies scoring at least some concerns or high overall risk of bias respectively (Figure 2).

**Figure 2:**
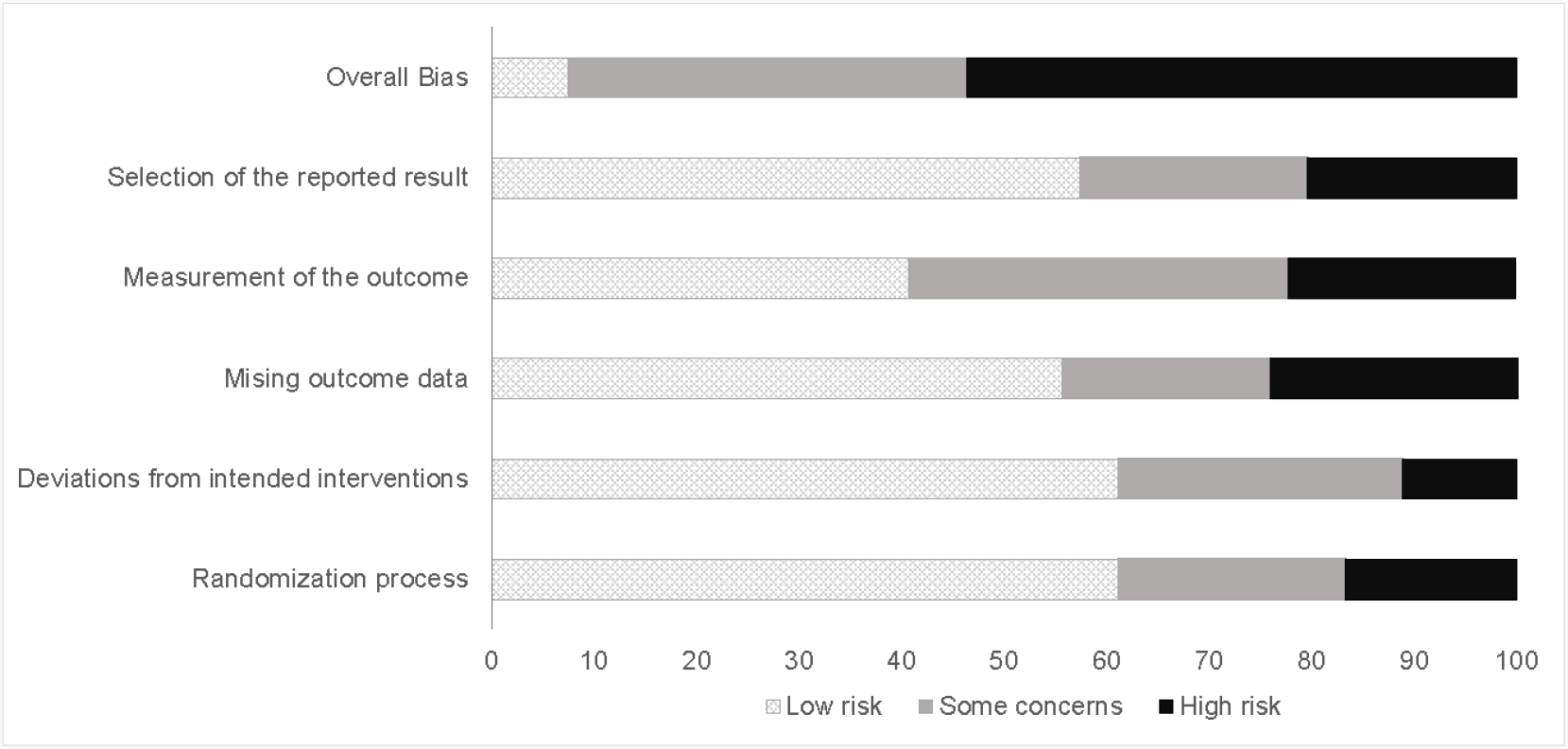
Domain level risk of bias summary and overall risk of bias across included studies.

### Patient demographics

The data extraction revealed 5091 patients who were treated subsequent to ACL rupture with a median age at treatment of 27.7 (IQR 25.7 - 30) with 32.6% (IQR 22.2 - 44.9) female. 4991 patients underwent ACLR; 100 patients were treated without surgical intervention (physiotherapy / rehabilitation). Height, weight and presence of comorbidities for included participants across studies were poorly reported. The surgical details extracted from the included articles varied for notchplasty, fixation methods, graft tissue, source, preparation, number and placement of tunnels and the approach (Table D).

**Table D:** Distribution of surgical technique characteristics within included papers and groups (N = 115) (%). *PT: Patella tendon: ST-only: Semitendinous-only; STG: Semitendinous and gracilis; TA: Tibialis anterior; Auto: Autograft; Allo: Allograft*.

| Notchplasty | Fixation | Graft Tissue | Graft Source | Graft preparation | Tunnel | Approach | Placement |
| --- | --- | --- | --- | --- | --- | --- | --- |
| Routine (15.6) | Combination (42.6) | PT (37.38) | Auto (86.1) | Bone-tendon (36.5) | Single (6870) | Anteromedial (35.7) | Anatomic-cent re (40.0) |
| As Required (3.5) | Screw (31.3) | ST-only (12.2) | Allo (7.8) | Quadruple (33.0) | Double (10.4) | Transtibial (26.1) | Anatomic - Other (12.2) |
| Never (4.4) | Suspension (9.5) | STG (409) | Not Reported (6.1) | Double (16.5) | Not Reported (20.9) | Rear (2.6) | Non anatomic/ Isometric (10.4) |
| Not Reported (76.5) | Not Reported (16.5) | TA(0.9) |  | Other (5.1) |  | Arthrotomy (3.5) | Not Reported (37.4) |
|  |  | Other (2.6) |  | Not Reported (9.7) |  | Not Reported (32.2) |  |
|  |  | Not Reported (4.4) |  |  |  |  |  |

### Outcomes

The included papers reported a median follow up from treatment of 4.9 months (IQR 1.9 - 24) across all groups and time points for included papers.The reporting of LOE incidence was inconsistent, with half the included studies requiring a t-norm conversion from knee angle summary statistics to an estimated incidence of LOE (%) at a cutoff threshold of 3 degrees. Studies that did define “full extension” of the knee to determine degree of extension loss used varied definitions **(Supplementary Material C)**. The method and conditions of measurement were also poorly reported (Table E), with a majority of studies defining full knee extension as relative to the contralateral limb or anatomical zero as determined by goniometry.. Knee extension measured with a goniometer, compared to the contralateral limb, with a 3 degree threshold to define a loss of extension, appeared to be the most common approach (Table E). Cutoff thresholds to determine the incidence of extension deficit (relative to full extension) also varied between studies.

**Table E:** Distribution of measurement characteristics (%).

| **Movement type** | **Position** | **Measurement Tool** | **Knee angle definition** | **Extension Loss Comparator** |
| --- | --- | --- | --- | --- |
| Active (9.4) | Straight leg - Prone (17.0) | Goniometer (60.4) | 3° (66.0) | Contralateral (73.6) |
| Passive (49.1) | Straight leg - Supine (37.7) | Manual (1.9) | 5° (34.0) | Anatomical zero (13.2) |
| Not Reported (41.5) | Straight leg - Seated (1.9) | Ruler (9.4) |  | Not Reported (13.2) |
|  | Not Reported (43.4) | Other (3.8) |  |  |
|  |  | Not Reported (24.5) |  |  |

#### Classification analysis

A sample of 276 timepoints with 40 fields collected on each timepoint were available for the classification analysis. The data was zero-inflated with a high proportion of loss of extension incidence (LOE) <5% (Figure 3). The reduced predictor set for classification component of analysis, after screening included the following variables: paper and group ID; time of follow up; average age at follow up; graft type; movement paradigm; sample size, graft placement/approach; randomisation factor; tissue type. The misclassification error for the model was found to be 16.7%, suggesting that the reduced predictor set provided a reasonable fit to the data. The model indicated that the study and observed group within the study were the most important factors for whether a timepoint was reported as undetected (incidence <1%) or otherwise, followed by time to follow up and graft type (Figure 4).

**Figure 3:**
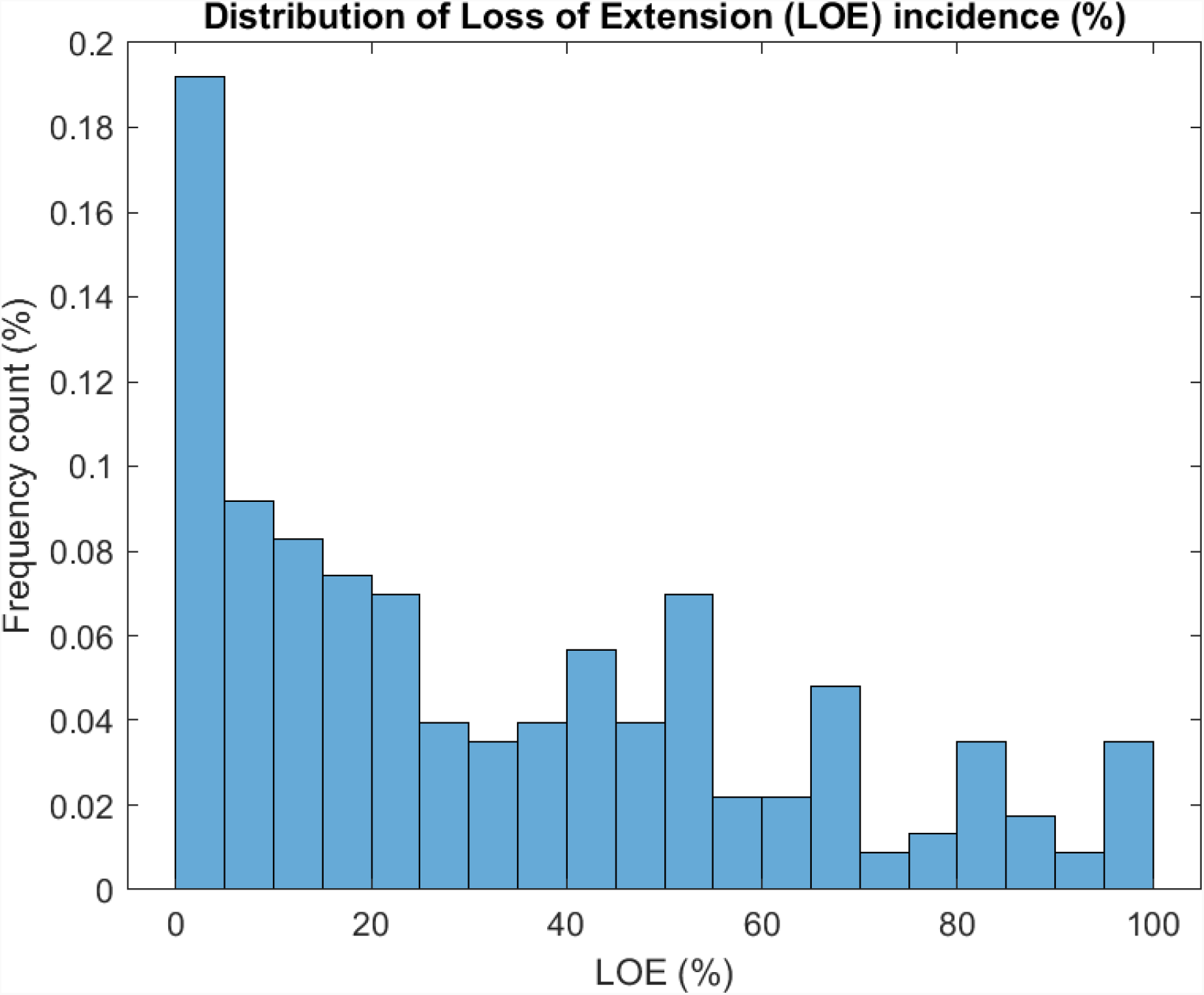
Distribution of LOE incidence from reported studies and timepoints extracted

**Figure 4:**
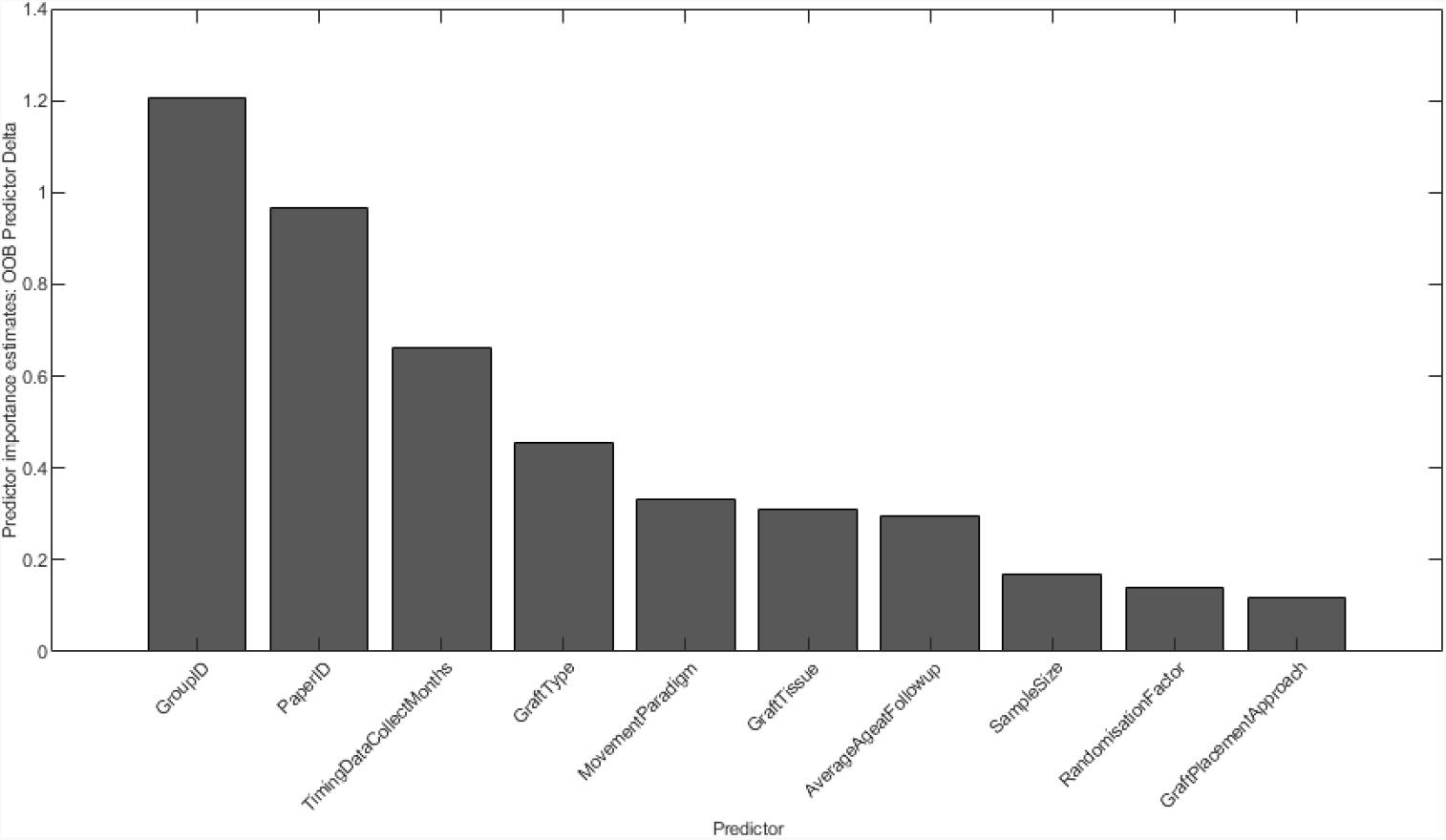
Estimation of predictor importance for reduced predictor set by permutation of out-of-bag (OOB) predictor observations.

#### Regression analysis

After subsetting the dataset for non-zero LOE, the generalised mixed effects linear model was performed on 229 observations from 43 eligible studies. The model provided a reasonable fit to the data (adjusted R^2^ = 35.2%) and revealed a significant negative association with time to follow up (Figure 5, Table F) and graft type on reported loss of extension incidence (%) (Table F; **Supplementary Material D - GLME Fixed**). However, some model miscalibration to the data was observed, with the upper confidence interval of the model intercept exceeding 100% (Table F). Nevertheless, the model indicated significant associations between Risk of Bias ratings, further analysis of *graft type* revealed lower LOE incidence for allograft and autograft compared to those papers that did not report a graft type used. However, the allograft data was derived from one study comparing allograft hamstring and patellar tendon grafts ^62^]. The model predictions for LOE relative to followup revealed an estimated incidence of 31.5% (95%CI 30.4 - 32.7) at 12months and 23.1% (95%CI 22.0 - 23.6%) at 24months (Figure 5 - bottom). The estimated average LOE incidence by time of followup can be estimated with Equation 2.

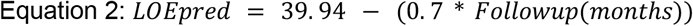

**Table F:** Summary of *significant fixed effects* from generalised linear mixed effects model.

| Predictor | Coefficient | UpperCI | LowerCI | P-Value |
| --- | --- | --- | --- | --- |
| Intercept | 84.1 | 50.9 | 117.3 | <0.001 |
| ROB (vs Some Concerns)<br>- High<br>- Low | -9.0<br>12.3 | -14.8<br>4.0 | -3.2<br>20.6 | 0.002<br>0.004 |
| Movement - NR vs Straight-Leg Supine | -12.0 | -21.3 | -2.7 | 0.012 |
| Followup (Months) | -0.43 | -0.75 | -0.12 | 0.008 |
| Graft Fixation - Screw | 13.3 | 2.5 | 24.0 | 0.016 |
| GraftType (vs NR)<br>- Allograft<br>- Autograft | -19.4<br>-13.8 | -34.2<br>-23.1 | -4.7<br>-4.4 | 0.01<br>0.004 |

\*NR - Not Reported

**Figure 5:**
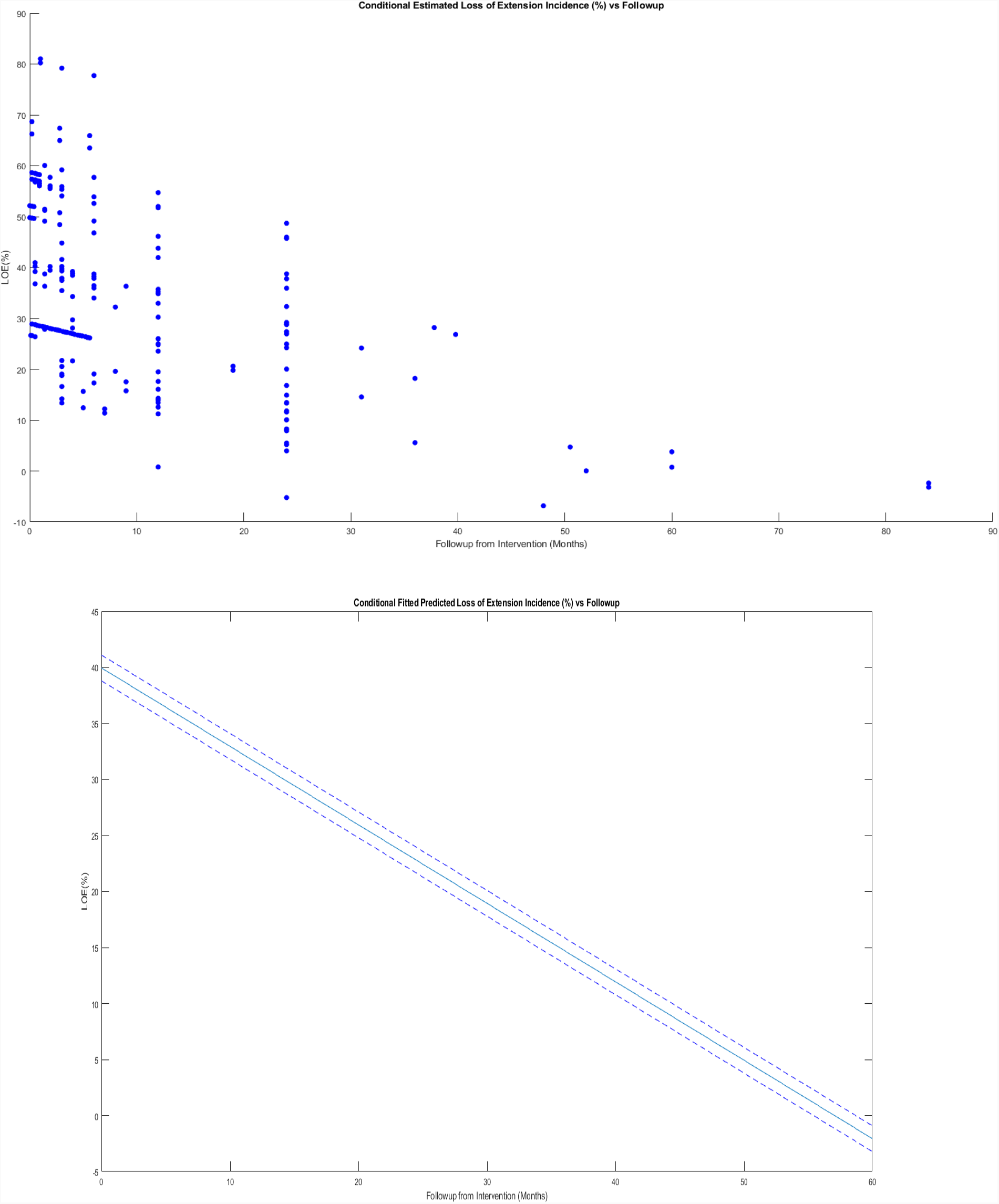
Relationship between follow up and predicted LOE from non-zero LOE reports (43 studies) (top) and linear fit of the conditional estimates of LOE with 95% confidence intervals (bottom)

## DISCUSSION

Restoring full extension after injury to the ACL and its subsequent reconstruction is an important milestone in progressive rehabilitation ^78–80^. An inability to achieve full extension, particularly under weight bearing conditions, alters the points of stress concentration between tibia and femur ^81^ and has implications for delayed return-to-sport, patellofemoral joint pain as well as the onset of post-injury osteoarthritis ^82,83^.

The primary objective of this review was to identify the incidence of loss of knee extension angle in patients diagnosed with ACL rupture electing to undergo formal treatment under the care of a registered clinical provider (including non-operative and arthroscopic ACL reconstruction) compared to the contralateral limb or control patients, as reported within randomised control studies. We report an overall average incidence of 31.5% (95%CI 30.4 - 32.7) at 12months. The secondary objective of this review was to explore the manner in which knee extension deficits have been reported and defined across the literature. We have noted substantial variability within the reporting of characteristics of LOE measurement across studies, including differences in the definition of a “normal” or “full” extension comparator for determining the degree of deficit, thresholds for categorising LOE and whether the ROM was determined via an active or passive movement paradigm (Table E). Methods for determining the knee extension were also varied, with some studies reporting extension deficits through measurement of heel height difference compared to the contralateral limb, in lieu of direct measurement of plane knee angles ^31,38,40,47,67^. Furthermore, some aspects of ROM measurements were not reported in up to 43% of papers reviewed (Table E) suggesting that a standardised framework for reporting measurement methods would be beneficial for a more accurate assessment of the incidence of LOE after ACL rupture and reconstruction.

Factors affecting the incidence of extension deficits were also examined in this systematic review. The general findings from the classification analysis revealed that the group in which the study participants were placed and studies themselves were the most important predictors for whether a time point was to detect an incidence of extension deficit or reported as undetected (incidence <1%). These results suggest that methodological differences in the papers contributed substantially to the reporting of LOE. In the majority of trials, LOE was reported as a secondary outcome, with definitions and measurement methods poorly reported. It is likely that at least some studies operationally defined LOE as symptomatic presentations, leading to selective measurement. This highlights the methodological limitations of the systematic review and potential influences of author bias, author background/specialties, interests and motivations, or regional / socioeconomic / cultural factors on the study design, and hence the detection of LOE within the cohorts of the included sample of studies. The predictor importance rankings for timing of data collection (or length of follow up), graft type and movement paradigm are further suggestive that the methodologies employed within the studies are also important determinants on reported LOE incidence.

The presence of LOE after ACLR is multifactorial ^7,84,85^, with graft impingement, arthrofibrosis, suboptimal graft placement and over-tensioning of the graft among the most common reported causes ^7,84^. While re-operation is occasionally required to treat severe extension deficits, cases where extension deficits resolve without further operative intervention and the cross-sectional findings of the present review suggest there may be an innate process by which ROM is recovered. The significant association between length of followup and LOE incidence identified by metaregression further supports this notion. We hypothesise that in these cases where knee extension recovers spontaneously, other processes such as soft tissue remodelling may be the primary mechanism. Of particular interest to note is the significant association between graft type used in ACL reconstruction on LOE incidence identified in this review, suggesting that graft type itself may contribute to the processes involved in extension recovery. However, this should be interpreted with caution due to limitations of the data analysis package which referenced the “not-reported” label for the graft type variable category, and limited number of datapoints available for analysis pertaining to studies reporting the use of allografts, which is further reflected by the wide confidence intervals observed around the fixed effect coefficient. While identifying the mechanism by which knee extension is recovered was outside the scope of this review, approximately 43% of included studies to this review were randomised based on graft type, indicating graft type remains an area of interest, and could guide future research objectives.

While the primary focus of this systematic review was to summarise the outcomes of interest within studies of randomised controlled design to improve the body of evidence, limitations stemming from poor standardisation and reporting of measurement methods, definition of knee angle and extension comparator across studies should be considered. The t-norm conversion to estimate the incidence of LOE for studies reporting knee angle or loss of extension angle may have possibly introduced errors within the meta-analysis due to its assumption of normal distribution for knee extension. When combined with the classification analysis and regression modelling, the methods described were considered most appropriate for determining overall incidence of extension deficits within the included sample of studies and for identifying factors that may be associated with the LOE incidence. However the influence of factors such as BMI or time to intervention were unable to be inferred due to the substantial rates of non-reporting. Further to this, substantial risks of bias were detected across a notable number of studies, with upwards of half the included studies scoring a high risk of bias in at least one domain, or multiple judgements of some concern across multiple domains. It may be noted that knee extension angle, the degree of extension deficit or incidence of LOE were often not the primary outcome of the study, rendering them less sensitive to detecting an incidence of LOE in the study and contributing to the biases detected with respect to the measurement, presentation and reporting of findings for these metrics. We note that the limited prioritisation of knee extension as an outcome in its own right within the included randomised trials is a likely contributor to the issues identified. Other factors suspected of influence over short-term postoperative knee extension, such as graft tensioning (tension magnitude and knee angle at which tensioning is achieved) and adjunct stabilisation procedures (e.g. lateral extra-articular tenodesis), may be better explored in non-randomised investigations. Further analysis is recommended on this component of the dataset collected as part of the current work.

## CONCLUSIONS

This review established the trajectory of knee extension incidence after ACL reconstruction in studies of RCT design. Evidence of the incidence and factors associated with loss of extension were identified; however knee extension after ACL reconstruction was measured at variable time points with a variety of measurement techniques across a mixed cohort composition, with poor descriptions of how full extension was standardised. Overall, up to 1 in 3 patients can be expected to present with loss of extension of at least 3 degrees at 12month followup, which may decrease to 1 in 4 at 2 years. Further work is required to better describe the natural history of LOE after ACL injury and reconstruction, with particular attention to the development of standardised methods and objective measurement within accepted movement paradigms.

## Supporting information

Supplementary material A: Study search strategy

Supplementary material B: Combine screening code

Supplementary material C: Summary of studies

Supplementary material D: GLME fixed

## Data Availability

Data is available on request.

## Acknowledgements

The authors would like to acknowledge the contribution of Binglong Lee, Mathew Holt, Michael Erian and Christopher Erian for their contribution to paper screening and data extraction, and Milad Ebraihimi for performing the data quality control.

## Protocol deviations

- The results for the proposed systematic review protocol ^14^ will be reported in a series of separate manuscripts categorised by study design and measurement methods. The methods and results presented herein describe the analysis of randomised control studies collected within the framework of the systematic review protocol.
- Embase via Ovid SP, Scopus and SPORTDiscus via EBSCO, AMED, CINAHL, LILACS, Scielo & Web of Knowledge were not directly searched as per the original protocol, based on the outcome of a pilot search conducted at the commencement of the study which indicated that the articles were indexed in Cochrane and Pubmed databases already.
- ROB2.0 was used to assess risk of bias to facilitate assessment of studies of RCT design.
- Alternative software was used to conduct data analysis.

