## Supplementary material A: Study search strategy for "Incidence and prognostic factors of knee extension deficits following anterior cruciate ligament reconstruction: A systematic review and meta-analysis of randomised controlled trials"

**All**
(ACL OR "anterior cruciate ligament") AND (knee OR tibiofem* OR femor* OR tibia* OR femur) NOT (adolesc* OR paediat* OR revis* OR immat* OR caus* OR arthroplast* OR "knee replacement" OR prosthes* OR prevent* OR cadaver* OR in-vitro OR osteoarthrit* OR arthrit* OR arthropath* OR gonarthro* OR "ankle instability" OR shoulder OR "posterior cruciate" OR kidney OR renal OR *lipid OR antibody)

**Treatment** (((((ACL OR "anterior cruciate ligament"))) AND ((rehab* OR inject* OR nonoperative OR conservative OR surgical OR surgery OR repair OR reconstruction OR *graft OR arthroscop*))) NOT ((adolesc* OR paediat* OR revis* OR immat* OR caus* OR arthroplast* OR replacement OR prosthes* OR prevent* OR cadaver* OR in-vitro OR osteoarthrit* OR arthrit* OR arthropath* OR gonarthro* OR "ankle instability" OR shoulder OR "posterior cruciate")))

**Clinical Outcomes** (((((ACL OR "anterior cruciate ligament"))) AND ((extension OR "fixed flexion" OR "loss of extension" OR "extension loss" OR stiffness OR arthrofibrosis OR imping* OR "minimum flexion" OR "range of motion" OR notch* OR cyclops)) NOT ((paediat* OR revis* OR immat* OR caus* OR arthroplast* OR replacement OR prosthes* OR prevent* OR cadaver* OR in-vitro OR osteoarthrit* OR arthrit* OR arthropath* OR gonarthro* OR "ankle instability" OR shoulder OR "posterior cruciate")))

**Biomechanics** (ACL OR "anterior cruciate ligament") AND (walk* OR jog* OR run* OR locomot* OR ambulat* OR stair* OR gait* OR stop* OR land* OR hop*) AND (kinemat* OR biomech* OR ang* OR motion OR kinet*) NOT (paediat* OR revis* OR immat* OR injury OR caus* OR arthroplast* OR replacement OR prosthes* OR prevent* OR cadaver* OR in-vitro OR osteoarthrit* OR arthrit* OR arthropath* OR gonarthro* OR "ankle instability" OR shoulder OR "posterior cruciate")
