## Supplementary material B: Combine screening code for "Incidence and prognostic factors of knee extension deficits following anterior cruciate ligament reconstruction: A systematic review and meta-analysis of randomised controlled trials"

### Code used to filter publication search results

```
#set required library
library(synthesisr)
#move to the correct input folder
setwd("G:/My Drive/EBMA/Client Drive/QEII Jubilee/EBMA Working/
Publications/ACLR Knee Ext SR Update_CB035Sep21/Data Collection")

# Retrieve file names in search results folder

bibfiles <- list.files('./Primary Search/',  full.names = TRUE)
print(bibfiles)

# Read in bibliographic files and store them in data.frame(s)
setwd("G:/My Drive/EBMA/Client Drive/QEII Jubilee/EBMA Working/
Publications/ACLR Knee Ext SR Update_CB035Sep21/Data Collection/Primary
Search")

paperpile_df <- read_refs(filename = 'Paperpile - ACLR SR Search Update Oct
2021 - Dec 08.ris',tag_naming = 'best_guess', verbose = TRUE, return_df =
TRUE)
cochrane_df1 <- read_refs(filename = 'Cochrane reviews-
export.ris',tag_naming = 'best_guess', verbose = TRUE, return_df = TRUE)
cochrane_df2 <- read_refs(filename = 'Cochrane trials-
export.ris',tag_naming = 'best_guess', verbose = TRUE, return_df = TRUE)
## dimensions_df <- read_refs(filename = 'Dimensions
BMKOAMSCSR.ris',tag_naming = 'best_guess', verbose = TRUE, return_df =
TRUE)
lens_df1 <- read_refs(filename = 'Lens - ACLLOEBiomech.ris',tag_naming =
'best_guess', verbose = TRUE, return_df = TRUE)
lens_df2 <- read_refs(filename = 'Lens - ACLLOEClinical.ris',tag_naming =
'best_guess', verbose = TRUE, return_df = TRUE)
lens_df3 <- read_refs(filename = 'Lens - ACLLOETreatment.ris',tag_naming =
'best_guess', verbose = TRUE, return_df = TRUE)
pubmed_df1 <- read_refs(filename = 'pubmed-ACLORanter-
clinical.nbib',tag_naming = 'best_guess', verbose = TRUE, return_df = TRUE)
pubmed_df2 <- read_refs(filename = 'pubmed-ACLORanter-
biomech.nbib',tag_naming = 'best_guess', verbose = TRUE, return_df = TRUE)
pubmed_df3 <- read_refs(filename = 'pubmed-ACLORanter-
treatment.nbib',tag_naming = 'best_guess', verbose = TRUE, return_df =
TRUE)

#Add in preprint search results
setwd("G:/My Drive/EBMA/Client Drive/QEII Jubilee/EBMA Working/
Publications/ACLR Knee Ext SR Update_CB035Sep21/Data Collection/Primary
Search - Grey")
load("preprintresults.RData")

#########################################################################################

setwd("G:/My Drive/EBMA/Client Drive/QEII Jubilee/EBMA Working/
Publications/ACLR Knee Ext SR Update_CB035Sep21/Data Collection")

## Line up headers
paperpilehead <- matrix(colnames(paperpile_df),dimnames = list(c(NULL),
c("Headers")))
cochranehead <- matrix(colnames(cochrane_df1),dimnames = list(c(NULL),
c("Headers")))
# dimensionshead <- matrix(colnames(dimensions_df),dimnames = list(c(NULL),
c("Headers")))
```

```
lenshead <- matrix(colnames(lens_df1),dimnames = list(c(NULL),
c("Headers")))
pubmedhead <- matrix(colnames(pubmed_df1),dimnames = list(c(NULL),
c("Headers")))
biorxivhead <- matrix(colnames(Result_biorxiv_df),dimnames = list(c(NULL),
c("Headers")))
medrxivhead <- matrix(colnames(Result_medrxiv_df),dimnames = list(c(NULL),
c("Headers")))

# Find out maximum length
max_ln <-
(max(length(paperpilehead),length(cochranehead),length(lenshead),length(pubmedhead),len

# Bring in headers into each column (fill remainder with NA) from https://
www.geeksforgeeks.org/create-dataframe-of-unequal-length-in-r/

InitialSet1 <- data.frame(col1 = c(paperpilehead,rep(NA, max_ln -
length(paperpilehead))),
           col2 = c(cochranehead,rep(NA, max_ln - length(cochranehead))),
           col3 = c(lenshead,rep(NA, max_ln - length(lenshead))),
           col4 = c(pubmedhead,rep(NA, max_ln - length(pubmedhead))),
           col5 = c(biorxivhead,rep(NA, max_ln - length(biorxivhead))),
           col6 = c(medrxivhead,rep(NA, max_ln - length(medrxivhead))))

names(InitialSet1)[1] <- "paperpile"
names(InitialSet1)[2] <- "cochrane"
# names(InitialSet1)[3] <- "dimensions"
names(InitialSet1)[3] <- "lens"
names(InitialSet1)[4] <- "pubmed"
names(InitialSet1)[5] <- "biorxiv"
names(InitialSet1)[6] <- "medrxiv"

##Have moved the InitialSet1 frame to project mastersheet [https://
docs.google.com/spreadsheets/d/1q6XnxdFTi7Usx7TE8hYq-
qKoPLXNbodmt5cv9JaKZx4/edit#gid=1338665864] to visualize each field

#DatabaseFields <-
c("title","year","journal","authors","abstract","doi","url","article
type","filename","keywords") #dont need to prespecify matrices in R

#Option 1: rename columns in each dataframe to common language and rbind to
append rows within next combined dataframe

library (data.table)
setnames(paperpile_df, old = c("source_type", "author","source"), new =
c("article_type","authors","journal"))
setnames(cochrane_df1, old = c("source_type", "A1","accession_zr"), new =
c("article_type","authors","database_id"))
setnames(cochrane_df2, old = c("source_type",
"A1","N2","concepts","accession_zr"), new =
c("article_type","authors","abstract","keywords","database_id"))
#setnames(dimensions_df, old = c("TY", "JO","author","PY","UR","DO","AN"),
new =
c("article_type","journal","authors","year","url","doi","database_id"))
setnames(lens_df1, old = c("source_type", "author","ID"), new =
c("article_type","authors","database_id"))
```

```
setnames(pubmed_df1, old =
c("publication_type","article_id","date_published", "author","pubmed_id"),
new = c("article_type","doi","year","authors","database_id"))
setnames(lens_df2, old = c("source_type", "author","ID"), new =
c("article_type","authors","database_id"))
setnames(pubmed_df2, old =
c("publication_type","article_id","date_published", "author","pubmed_id"),
new = c("article_type","doi","year","authors","database_id"))
setnames(lens_df3, old = c("source_type", "author","ID"), new =
c("article_type","authors","database_id"))
setnames(pubmed_df3, old =
c("publication_type","article_id","date_published", "author","pubmed_id"),
new = c("article_type","doi","year","authors","database_id"))
setnames(Result_biorxiv_df, old = c("ID","date","link_page"), new =
c("database_id","year","url"))
setnames(Result_medrxiv_df, old = c("ID","date","link_page"), new =
c("database_id","year","url"))

######Refactor publication year for each input dataframe###

library(stringi)

paperpile_df$year2 <- stri_match_first_regex(paperpile_df$year,"\\D*(\
\d{4})")
cochrane_df1$year2 <- stri_match_first_regex(cochrane_df1$year,"\\D*(\
\d{4})")
cochrane_df2$year2 <- stri_match_first_regex(cochrane_df2$year,"\\D*(\
\d{4})")
lens_df1$year2 <- stri_match_first_regex(lens_df1$year,"\\D*(\\d{4})")
lens_df2$year2 <- stri_match_first_regex(lens_df2$year,"\\D*(\\d{4})")
lens_df3$year2 <- stri_match_first_regex(lens_df3$year,"\\D*(\\d{4})")
Result_biorxiv_df$year2 <- stri_match_first_regex(Result_biorxiv_df$year,"\
\D*(\\d{4})")
Result_medrxiv_df$year2 <- stri_match_first_regex(Result_medrxiv_df$year,"\
\D*(\\d{4})")
pubmed_df1$year2 <- stri_match_first_regex(pubmed_df1$year,"\\D*(\\d{4})")
pubmed_df2$year2 <- stri_match_first_regex(pubmed_df2$year,"\\D*(\\d{4})")
pubmed_df3$year2 <- stri_match_first_regex(pubmed_df3$year,"\\D*(\\d{4})")


#Last dataframe before screening; include year2 (refactored to a year)
instead of original year field
library(tidyverse)
paperpile_reduce <- paperpile_df %>%
select(article_type,year2,authors,title,abstract,journal,doi,url,notes)
#missing database_id; added notes
cochrane_reduce1 <- cochrane_df1 %>%
select(article_type,year2,authors,title,abstract,journal,doi,url,database_id)
#dimensions_reduce <- dimensions_df1 %>%
select(article_type,year,authors,title,journal,doi,url,database_id)
#missing abstract
lens_reduce1 <- lens_df1 %>%
select(article_type,year2,authors,title,abstract,journal,doi,url,database_id)
pubmed_reduce1 <- pubmed_df1 %>%
select(article_type,year2,authors,title,abstract,journal,doi,database_id)
#missing url
```

```
cochrane_reduce2 <- cochrane_df2 %>%
select(article_type,year2,authors,title,abstract,journal,doi,url,database_id)
#dimensions_reduce <- dimensions_df1 %>%
select(article_type,year,authors,title,journal,doi,url,database_id)
#missing abstract
lens_reduce2 <- lens_df2 %>%
select(article_type,year2,authors,title,abstract,journal,doi,url,database_id)
pubmed_reduce2 <- pubmed_df2 %>%
select(article_type,year2,authors,title,abstract,journal,doi,database_id)
#missing url

lens_reduce3 <- lens_df3 %>%
select(article_type,year2,authors,title,abstract,journal,doi,url,database_id)
pubmed_reduce3 <- pubmed_df3 %>%
select(article_type,year2,authors,title,abstract,journal,doi,database_id)
#missing url

biorxiv_reduce <- Result_biorxiv_df %>%
select(year2,authors,title,abstract,doi,database_id,published) #missing
article_type,journal; added publication
medrxiv_reduce <- Result_medrxiv_df %>%
select(year2,authors,title,abstract,doi,database_id,published) #missing
article_type,journal; added publication

library(expss)
InitialSet2 =
add_rows(paperpile_reduce,cochrane_reduce1,cochrane_reduce2,lens_reduce1,lens_reduce2,l
= "add")

### Filter InitialSet2 by publication year
library(dplyr)
InitialSet3 <- filter(InitialSet2, InitialSet2$year2 > 2019)

#########################################################################################
#De-duplicate titles
# Remove articles that have identical titles
InitialSet4 <- deduplicate(InitialSet3,match_by = "title",method = "exact")

# Remove articles that have identical doi
InitialSet5 <- deduplicate(InitialSet4,match_by = "doi",method = "exact")



library(revtools)
library(tictoc)
# Use string distance to identify likely duplicates
tic("String Distance Deduplicate")
duplicates_string <- find_duplicates(InitialSet5,"title",method =
"osa",to_lower = TRUE,threshold = 7)
toc()

# we can extract the line numbers from the dataset that are likely
duplicated
# this lets us manually review those titles to confirm they are duplicates

manual_checks <- review_duplicates(InitialSet5[[3]], duplicates_string)
print(manual_checks)
```

```
# now we can extract unique references from our dataset
# we need to pass it the dataset (df) and the matching articles
(new_duplicates)
Combine_Screen <- extract_unique_references(InitialSet5,duplicates_string)

## screen for reviews and meta-analyses

## Filter for reviews/SR/MA etc

ArticleTypes = c("systematic review", "meta-analysis", "meta analysis",
"metaregression")

#https://stackoverflow.com/questions/38940847/filter-a-column-based-on-a-
list-of-words-in-r
#loop through ArticleTypes and perform regex on the title column of results
dataframe
Review_Results <- Combine_Screen[Reduce(`|`, lapply(ArticleTypes, grepl, x
= Combine_Screen$title)),]
length(Review_Results$title)

#write.csv(review_results, file = "reviewresults.csv")

#Export as CSV for import into Rayyan
write.csv(Combine_Screen, file = "CombineScreen.csv")

## Filter for RCTs
Inclusions = c("randomised controlled trial", "randomised", "randomized
controlled trial", "randomized","clinicaltrials.gov","ICRTP")

#loop through ArticleTypes and perform regex on the title column of results
dataframe
RCT_Results <- Combine_Screen[Reduce(`|`, lapply(Inclusions, grepl, x =
Combine_Screen$title)),]
length(RCT_Results$title)
RCT_Results2 <- Combine_Screen[Reduce(`|`, lapply(Inclusions, grepl, x =
Combine_Screen$abstract)),]
length(RCT_Results2$title)

# Save as rdata files to read back in
setwd("G:/My Drive/EBMA/Client Drive/QEII Jubilee/EBMA Working/
Publications/ACLR Knee Ext SR Update_CB035Sep21/Data Collection/Combine
Screen")
save (Combine_Screen,Review_Results,RCT_Results2, file =
"CombineScreenMay2022.RData")

write.csv(RCT_Results2, file = "RCT_ResultsMay2022.csv")

# Save all objects in Object.RData
save.image(file = "CombineScreenMay2022.RData")
```

**Code used to filter grey literature search results**

```
#Load required libraries
library(medrxivr)

#move to the correct input folder
setwd("G:/My Drive/EBMA/Client Drive/Resources/Preprint Databases")

## Load up biorxiv and medrxiv database files as of: 8-Nov-2021

load('Preprint_Bio.Rdata')
load('Preprint_Med.Rdata')

#move to the correct input folder
setwd("G:/My Drive/EBMA/Client Drive/Resources/Clinical Trials/
ICTRPWeek1November2021")

#library(tidyverse)

## Load up ICRTP (WHO) database files as of: 4-Nov-2021
ICRTP <- read.csv("ICTRPWeek1November2021.csv")

#####################################################################################Se

# Perform an advanced search
topic1  <- c("anterior cruciate ligament","ACL")  # Combined with Boolean
OR
# topic2  <- c(""."","")               # Combined with Boolean OR
myquery <- list(topic1, topic2)                            # Combined with
Boolean AND

Result_medrxiv <- mx_search(data = preprint_med, query = topic1)
Result_biorxiv <- mx_search(data = preprint_bio, query = topic1)

#https://stackoverflow.com/questions/38940847/filter-a-column-based-on-a-
list-of-words-in-r
#loop through ArticleTypes and perform regex on the title column of results
dataframe
Result_ICRTP <- ICRTP[Reduce(`|`, lapply(topic1, grepl, x =
ICRTP$Scientific_title)),]
print(length(Result_ICRTP$Scientific_title))


######################################################################################E
#Export results to be combined with search results from published
literature

#move to the correct output folder
setwd("G:/My Drive/EBMA/Client Drive/QEII Jubilee/EBMA Working/
Publications/ACLR Knee Ext SR Update_CB035Sep21/Data Collection/Primary
Search - Grey")

# Save as rdata files to read back in and combine with search from
published material
Result_biorxiv_df <- as.data.frame(Result_biorxiv)
Result_medrxiv_df <- as.data.frame(Result_medrxiv)
save (Result_biorxiv_df,Result_medrxiv_df, file = "preprintresults.RData")

#library(synthesisr)
```

```
#write_refs(as.data.frame(Result_medrxiv),format = "ris", file =
"Results_medrxiv")
#write_refs(as.data.frame(Result_biorxiv),format = "ris", file =
"Results_biorxiv")
##File size is likely reduced due to lack of abstracts for trial
registration info
#write_refs(Result_ICRTP,format = "ris", file = "Results_ICRTP")
```
