## Supplementary material C: Summary of studies for "Incidence and prognostic factors of knee extension deficits following anterior cruciate ligament reconstruction: A systematic review and meta-analysis of randomised controlled trials"

| First Author | Year | Randomisation Method | Extension Comparator | Knee angle definition | Which outcomes were reported* | Overall RoB Rating |
|---|---|---|---|---|---|---|
| Noyes | 1987 | ROM restriction | Anatomical zero | Degrees short of full extension/zero | Knee extension angle (deg) | High |
| Richmond | 1991 | ROM restriction | Anatomical zero | Degrees short of full extension/zero | Knee extension angle (deg) | Some concerns |
| Engström | 1995 | ROM restriction | Contralateral | Not reported | Loss of knee extension (deg) | Some concerns |
| Feller | 1997 | ROM restriction | Contralateral | Not reported | Loss of knee extension (deg) | High |
| Zätterström | 1998 | Post rehab | Contralateral | Not reported | Loss of knee extension (deg) | High |
| Risberg | 1999 | ROM restriction | Not reported | Not reported | Loss of knee extension (deg) | Some concerns |
| Eriksson | 2001 | Graft type | Contralateral | Not reported | Loss of knee extension (%) | Some concerns |
| Feller | 2001 | Graft type | Contralateral | Heel Height Difference | Loss of knee extension (deg) | Some concerns |
| Möller | 2001 | ROM restriction | Anatomical zero | Degrees short of full extension/zero | Loss of knee extension (deg) | Some concerns |
| Anderson | 2002 | Graft type | Contralateral | Not reported | Loss of knee extension (%) | High |
| Henriksson | 2002 | ROM restriction | Contralateral | Relative to contralateral | Loss of knee extension (deg) | High |
| Shaieb | 2002 | Graft type | Not reported | Not reported | Loss of knee extension (%) | High |
| Feller | 2003 | Graft type | Contralateral | Not reported | Loss of knee extension (deg) | Some concerns |
| Melegati | 2003 | ROM restriction | Contralateral | Heel Height Difference | Loss of knee extension (%) | High |
| Mikkelsen | 2003 | ROM restriction | Anatomical zero | Degrees short of full extension/zero | Loss of knee extension (%) | High |
| Aglietti | 2004 | Graft type | Contralateral | Heel Height Difference | Loss of knee extension (%) | High |
| McDevitt | 2004 | ROM restriction | Contralateral | Not reported | Loss of knee extension (%) | Some concerns |
| Ibrahim | 2005 | Graft type | Contralateral | Not reported | Loss of knee extension (%) | High |
| Grant | 2005 | Post rehab | Contralateral | Relative to contralateral | Loss of knee extension (%) | Some concerns |
| Harilainen | 2005 | Graft fixation | Contralateral | Not reported | Loss of knee extension (%) | Some concerns |
| Isberg | 2006 | ROM restriction | Contralateral | Degrees short of full extension/zero | Knee extension angle (deg) | Some concerns |
| Heijne | 2007 | Graft type | Contralateral | Not reported | Loss of knee extension (%) | High |
| Järvelä | 2007 | Graft type | Contralateral | Not reported | Loss of knee extension (%) | High |
| Lidén | 2007 | Graft type | Contralateral | Degrees short of full extension/zero | Multiple | Low |
| Maletis | 2007 | Graft type | Contralateral | Heel Height Difference | Loss of knee extension (%) | Some concerns |
| Bottoni | 2008 | Surgical timing | Contralateral | Not reported | Loss of knee extension (deg) | High |
| Siebold | 2008 | Graft type | Contralateral | Not reported | Loss of knee extension (deg) | Low |

| | | | | | | |
|---|---|---|---|---|---|---|
| Mayr | 2010 | ROM restriction | Contralateral | Not reported | Loss of knee extension (deg) | Some concerns |
| Grant | 2010 | Post rehab | Contralateral | Degrees short of full extension/zero | Loss of knee extension (deg) | High |
| Raviraj | 2010 | Surgical timing | Contralateral | Degrees short of full extension/zero | Loss of knee extension (deg) | Some concerns |
| Hussein | 2011 | Graft type | Not reported | Not reported | Loss of knee extension (deg) | Some concerns |
| Noh | 2011 | Graft type | Contralateral | Not reported | Loss of knee extension (%) | High |
| Cappellino | 2012 | Post rehab | Anatomical zero | Degrees short of full extension/zero | Knee extension angle (deg) | Some concerns |
| Hong | 2012 | ACL remnant | Not reported | Not reported | Knee extension angle (deg) | High |
| Ahldén | 2013 | Graft type | Contralateral | Degrees short of full extension/zero | Multiple | High |
| Christensen | 2013 | Post rehab | Contralateral | Not reported | Loss of knee extension (deg) | Some concerns |
| Zhu | 2013 | Post rehab | Contralateral | Not reported | Loss of knee extension (deg) | High |
| Dai | 2015 | Graft type | Not reported | Not reported | Loss of knee extension (deg) | High |
| Kang | 2015 | Graft type | Not reported | Not reported | Loss of knee extension (%) | Some concerns |
| Koga | 2015a | Graft placement | Contralateral | Not reported | Loss of knee extension (deg) | Some concerns |
| Koga | 2015b | Graft type | Contralateral | Not reported | Multiple | High |
| Mohtadi | 2015 | Graft type | Contralateral | Not reported | Loss of knee extension (%) | Some concerns |
| Mei | 2016 | Graft type | Contralateral | Degrees short of full extension/zero | Knee extension angle (deg) | High |
| Webster | 2016 | Graft type | Contralateral | Heel Height Difference | Loss of knee extension (deg) | High |
| Mousavi | 2017 | Graft fixation | Anatomical zero | Degrees short of full extension/zero | Loss of knee extension (%) | High |
| Eriksson | 2018 | Surgical timing | Contralateral | Degrees short of full extension/zero | Multiple | Low |
| Mohtadi | 2019 | Graft type | Contralateral | Not reported | Loss of knee extension (%) | Some concerns |
| Shumborski | 2019 | Graft fixation | Contralateral | Not reported | Loss of knee extension (%) | High |
| Getgood | 2020 | Adjunct procedure | Anatomical zero | Degrees short of full extension/zero | Knee extension angle (deg) | Low |
| Sonnery-Cottet | 2020 | Graft type; adjunct procedure | Contralateral | Degrees short of full extension/zero | Loss of knee extension (%) | High |
| von Essen | 2020 | Surgical timing | Contralateral | Degrees short of full extension/zero | Loss of knee extension (deg) | High |
| Asif | 2021 | Graft fixation | Anatomical zero | Degrees short of full extension/zero | Loss of knee extension (%) | High |
| Hamido | 2021 | Graft type | Anatomic zero | Degrees short of full extension/zero | Loss of knee extension (%) | High |
| Irrgang | 2021 | Graft type | Contralateral | Relative to contralateral | Multiple | Some concerns |

**N.B. Knee extension (deg) denotes that either the knee flexion/extension angle, extension deficit or heel height difference was reported in the study - extracted data may have been re-coded where necessary to represent a loss of extension where full extension was defined at zero degrees of flexion. Heel height differences were converted to degrees using previously described methods (Sachs et al. 1989).*
