## Supplementary material D: GLME fixed for "Incidence and prognostic factors of knee extension deficits following anterior cruciate ligament reconstruction: A systematic review and meta-analysis of randomised controlled trials"

| Name | Estimate | SE | tStat | DF | pValue | Lower | Upper |
|---|---|---|---|---|---|---|---|
| (Intercept) | 84.10 | 16.82 | 5.00 | 208.00 | 0.00 | 50.95 | 117.26 |
| ROBOverall_High | -8.99 | 2.95 | -3.05 | 208.00 | 0.00 | -14.81 | -3.18 |
| ROBOverall_Low | 12.28 | 4.22 | 2.91 | 208.00 | 0.00 | 3.96 | 20.60 |
| ExtensionThreshold_3 degrees | 2.96 | 2.56 | 1.16 | 208.00 | 0.25 | -2.09 | 8.01 |
| MovementParadigm2_ Not reported | -12.00 | 4.72 | -2.54 | 208.00 | 0.01 | -21.31 | -2.68 |
| MovementParadigm2_ Straight leg - prone | 1.98 | 4.94 | 0.40 | 208.00 | 0.69 | -7.76 | 11.72 |
| MovementParadigm2_ Straight leg - seated | 12.72 | 10.36 | 1.23 | 208.00 | 0.22 | -7.72 | 33.15 |
| TimingDataCollectMonths | -0.43 | 0.16 | -2.69 | 208.00 | 0.01 | -0.75 | -0.12 |
| AverageAgeatFollowup | -0.81 | 0.59 | -1.36 | 208.00 | 0.17 | -1.97 | 0.36 |
| Meniscal_CartilageInjuries_No | 2.76 | 6.91 | 0.40 | 208.00 | 0.69 | -10.86 | 16.37 |
| Meniscal_CartilageInjuries_Not Reported | 14.48 | 9.62 | 1.50 | 208.00 | 0.13 | -4.50 | 33.45 |
| Fixation_Combination | 3.66 | 5.27 | 0.70 | 208.00 | 0.49 | -6.72 | 14.05 |
| Fixation_Not Reported | -10.84 | 8.86 | -1.22 | 208.00 | 0.22 | -28.32 | 6.63 |
| Fixation_Screw | 13.28 | 5.45 | 2.44 | 208.00 | 0.02 | 2.55 | 24.02 |
| Fixation_Suspension | -8.57 | 6.81 | -1.26 | 208.00 | 0.21 | -21.99 | 4.85 |
| GraftType_Allo | -19.45 | 7.46 | -2.61 | 208.00 | 0.01 | -34.15 | -4.74 |
| GraftType_Auto | -13.76 | 4.74 | -2.90 | 208.00 | 0.00 | -23.11 | -4.42 |
| GraftPlacementApproach_AM | 3.87 | 4.62 | 0.84 | 208.00 | 0.40 | -5.25 | 12.99 |
| GraftPlacementApproach_Arthrotomy | -3.38 | 6.95 | -0.49 | 208.00 | 0.63 | -17.09 | 10.33 |
| GraftPlacementApproach_Not Reported | 6.39 | 6.08 | 1.05 | 208.00 | 0.29 | -5.60 | 18.38 |
| GraftPlacementApproach_Rear-entry | -5.52 | 12.04 | -0.46 | 208.00 | 0.65 | -29.26 | 18.21 |
